# Antibody response patterns in COVID-19 patients with different levels of disease severity—Japan

**DOI:** 10.1101/2020.11.20.20231696

**Authors:** Kazuo Imai, Yutaro Kitagawa, Sakiko Tabata, Katsumi Kubota, Mayu Nagura-Ikeda, Masaru Matsuoka, Kazuyasu Miyoshi, Jun Sakai, Noriomi Ishibashi, Norihito Tarumoto, Shinichi Takeuchi, Toshimitsu Ito, Shigefumi Maesaki, Kaku Tamura, Takuya Maeda

## Abstract

**Background:** We analyzed antibody response patterns according to level of disease severity in patients with novel coronavirus disease 2019 (COVID-19) in Japan.

**Methods:** We analyzed 611 serum specimens from 231 patients with COVID-19 (mild, 170; severe, 31; critical, 30). IgM and IgG antibodies against nucleocapsid protein (N) and spike 1 protein (S1) were detected by enzyme-linked immunosorbent assays.

**Findings:** The peaks of fitting curves for the OD values of IgM and IgG antibodies against N appeared simultaneously, while those against S1 were delayed compared with N. The OD values of IgM against N and IgG against both N and S1 were significantly higher in the severe and critical cases than in the mild cases at 11 days after symptom onset. The seroconversion rates of IgG were higher than those of IgM against both N and S1 during the clinical course based on the optimal cut-off values defined in this study. The seroconversion rates of IgG and IgM against N and S1 were higher in the severe and critical cases than in the mild cases.

**Conclusion:** Our findings show that a stronger antibody response occurred in COVID-19 patients with greater disease severity and there were low seroconversion rates of antibodies against N and S1 in the mild cases. The antibody response patterns in our population suggest a second infection pattern, leading us to hypothesize that cross-reactivity occurs between SARS-CoV-2 and past infection with other human coronaviruses.

## Introduction

Novel coronavirus disease 2019 (COVID-19), which is caused by severe acute respiratory syndrome coronavirus 2 (SARS-CoV-2) infection, was initially reported in December 2019 in Wuhan, China, [1] and it has since become an ongoing pandemic worldwide [2].

Patients with COVID-19 are predominantly asymptomatic or have mild symptoms, but approximately 20% of patients develop severe disease [3]. The worldwide scientific community is still searching for the mechanism of disease pathogenesis to identify an effective treatment. Recently, several reports have suggested that the antibody response against SARS-CoV-2 may be associated with disease severity [4-10]. Identifying the antibody response patterns of target populations and the differences in these patterns between patients according to disease severity will help to clarify the pathogenicity and humoral immunity for SARS-CoV-2. The main antigens of SARS-CoV-2 are the internal nucleocapsid protein (N) and external spike protein (S), which consists of 2 subunits (S1 and S2). In particular, S1 contains a receptor-binding domain (RBD) that is responsible for binding to the angiotensin-converting enzyme 2 receptor on host cells at the initiation of infection [11]; thus, antibodies targeting S1 and RBD are expected to inhibit angiotensin-converting enzyme 2/RBD binding and have neutralization activity [8, 12-15].

Differences in antibody response patterns against each antigen between patients with different levels of disease severity have been reported based on limited samples collected from specific countries [9, 12]; however, it is unclear whether these response patterns can be similarly applied to different populations. Here, we describe the differences of the antibody response patterns for N and S1 and isotypes among 611 serum specimens collected from 231 Japanese patients with COVID-19 with different levels of disease severity.

## Methods

### Patients with COVID-19 and serum specimens

A total of 611 serum specimens were analyzed from 231 patients with laboratory-confirmed COVID-19 who were referred to and hospitalized at Saitama Medical University Hospital and Self-Defense Forces Central Hospital in Japan from February 11 to May 23, 2020. Briefly, the patients’ age ranged from 18 to 93 years (median, 49 years; interquartile range [IQR], 38–66 years), and 138 patients (59.7%) were male and 93 (40.3%) were female. According to their presentation during hospitalization, the symptomatic cases were subdivided into 3 groups at the end of hospitalization. Severe symptomatic cases were defined as patients showing clinical symptoms of pneumonia (percutaneous oxygen saturation < 93% and need for oxygen therapy). Critical cases were defined as showing a need for oxygen therapy using a high-flow nasal cannula and non-invasive positive pressure ventilation or invasive mechanical ventilation. The remaining symptomatic cases were classified as mild cases. Among the 231 patients, 170 (74.0%), 31 (13.4%), and 30 (13.0%) were classified as having mild, severe, and critical COVID-19 at the end of the hospitalization period, respectively. All patients were examined for SARS-CoV-2 by quantitative reverse-transcription polymerase chain reaction (RT-qPCR) using pharyngeal and nasopharyngeal swabs collected at public health institutes and hospitals in accordance with the nationally recommended method in Japan [16]. In brief, the gene encoding the N protein of SARS-CoV-2 was amplified by RT-qPCR using the following sets of primers and probes. N-1 set: N_Sarbeco_F1, N_Sarbeco_R1, and N_Sarbeco_P1; N-2 set: NIID_2019-nCOV_N_F2, NIID_2019-nCOV_N_R2, and NIID_2019-nCOV_N_P2 [16]. Serum samples were collected on admission and during hospitalization. Briefly, the median number of collected serum specimens for all 231 patients was 2 (IQR, 1–3) samples, with 2 (IQR, 1–2) samples for the 170 mild cases, 3 (IQR, 2–4) samples for the 31 severe cases, and 3 samples (IQR, 2–6) for the 30 critical cases. All serum samples were stored at -80°C before use in enzyme-linked immunosorbent assays (ELISAs).

### Negative samples from patients without COVID-19

To determine the optimal cutoff value for each ELISA, we used 150 serum samples collected from 150 patients at Saitama Medical University Hospital, Japan, from April to October 2019, before SARS-CoV-2 was first reported in China. All serum samples were stored at -80°C before use in ELISAs.

### Detection of antibodies against SARS-CoV-2 by ELISAs

To measure antibody titers against N and S1, a QuaResearch COVID-19 Human IgM IgG ELISA Kit (Nucleocapsid Protein) (RCOEL961N; Cellspect Co., Ltd., Iwate, Japan) and QuaResearch COVID-19 Human IgM IgG ELISA kit (Spike Protein-S1) (RCOEL961S1; Cellspect Co., Ltd.) were used, respectively. These kits are based on the indirect ELISA method, and each kit comes with different immobilized antigenic proteins. The plate of the COVID-19 Human IgM IgG ELISA Kit (Nucleocapsid Protein) contains immobilized recombinant N protein (1–419 AA) of SARS-CoV-2 expressed in *Escherichia coli*. The plate of the COVID-19 Human IgM IgG ELISA Kit (Spike Protein-S1) contains immobilized recombinant S1 protein (S1, 251–660 AA) of SARS-CoV-2 expressed in *E. coli*. Serum and plasma samples were diluted 1:1000 in 1% bovine serum albumin/phosphate-buffered saline with Tween 20 (PBST) for ELISAs with N and S1 proteins. The plates were read at 450 nm with an automated ELISA system (QRC5LB925; Cellspect Co., Ltd.) in accordance with the manufacturer’s measurement protocol.

### Definitions

The timing of seroconversion was defined as when the serum specimen showed an optical density (OD) value for each ELISA above the determined cutoff OD value. The day of symptom onset was defined as day 1.

### Ethics statement

This study was reviewed and approved by the Institutional Review Board of Saitama Medical University (approval number 1917), Institutional Review Board of Saitama Medical University Hospital (approval numbers 20064.01 and 20001), and Institutional Review Board of the Self-Defense Forces Central Hospital (approval number 01-011).

### Statistical analysis

All serum samples were evaluated by ELISA in triplicate and the average OD value for these measurements was defined as the test result. Continuous variables were expressed as the mean and standard deviation (SD) or median and IQR, and compared using the *t*-test or Wilcoxon rank-sum test for parametric or non-parametric data, respectively. The optimum cutoff OD value for each ELISA was determined to minimize the OD value obtained from 150 negative samples from patients without COVID-19 to ensure specificity > 98.0%. All statistical analyses were conducted using R (v 4.0.2; R Foundation for Statistical Computing, Vienna, Austria; http://www.R-project.org/).

## Results

### Kinetics of the antibody response according to disease severity

The kinetics of the antibody response against each antigen according to disease severity are shown in **Figure 1**. For IgM-N, low peaks of the fitting curves were observed in the severe and critical cases, but not in the mild cases (**Figure 1A**). The peaks were observed approximately 15 days after symptom onset in the severe and critical cases (**Figure 1A**). The fitting curves for IgM-S1 stayed at a low level in all 3 groups (**Figure 1B**). The peaks of the fitting curves for IgG-N were observed approximately 18 days after symptom onset in the severe and critical cases and 25 days in the mild cases. The peaks were higher in the severe and critical cases than in the mild cases. The peaks of the fitting curves for IgG-S1 were delayed compared with IgG-N (**Figure 1C and 1D**); the peaks were observed approximately 28 days after symptom onset in the mild and critical cases and 40 days in the severe cases. The peak values were higher in the order of critical, severe, and mild cases **(Figure 1D)**.

**Figure 1.**
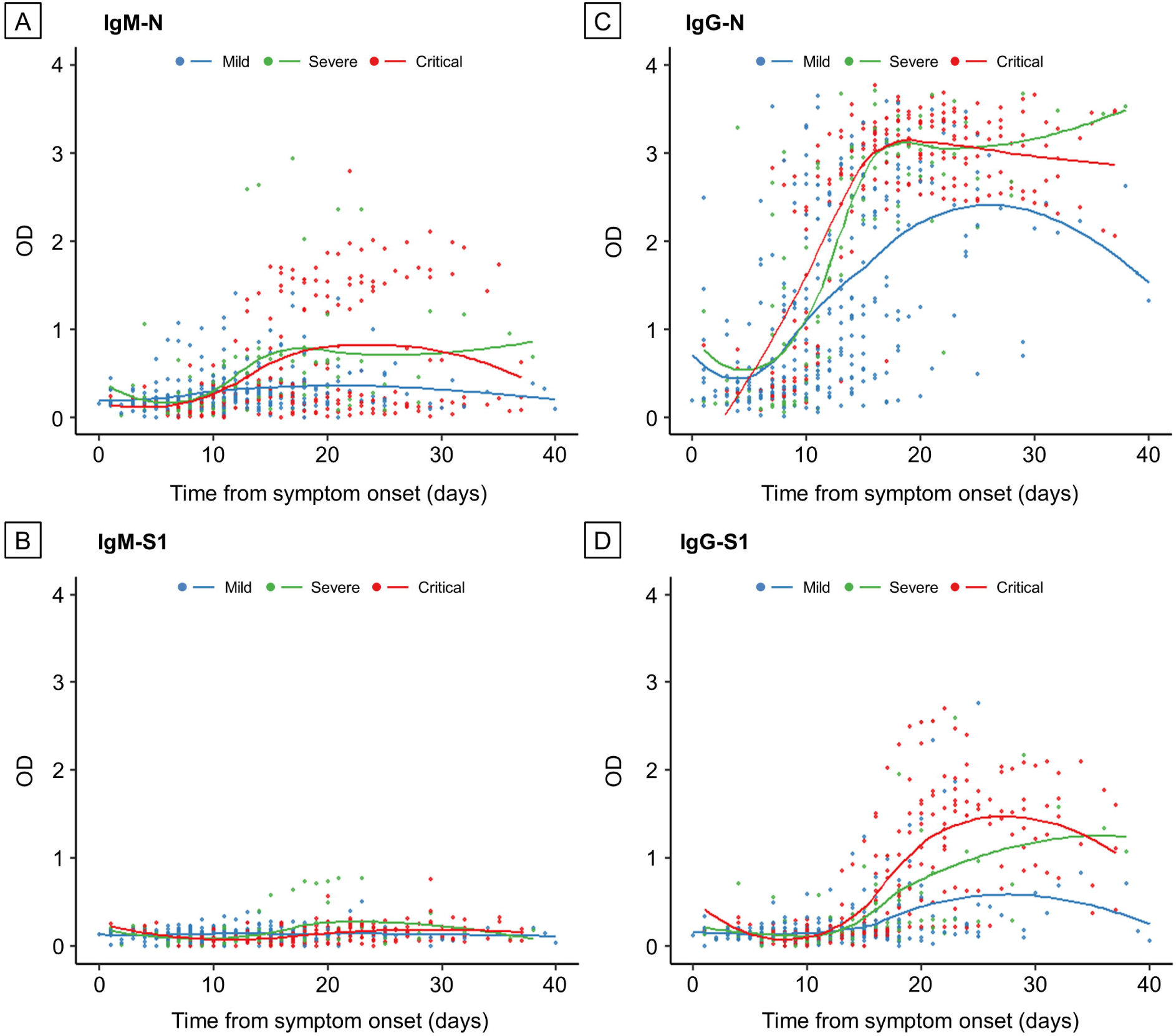
Kinetics of the IgM and IgG antibody responses according to disease severity. (A) IgM-N, (B) IgM-S1, (C) IgG-N, and (D) IgG-S1. Plots show time to sample collection from symptom onset and OD values for ELISAs. Blue plots and line, mild cases; green plots and line, severe cases; and red plots and line, critical cases.

Within 10 days after symptom onset, the OD values for IgM-N were lower in the critical cases than in the mild cases, and then the OD values increased rapidly in the critical cases (critical vs. mild, *p* = 0.017) (**Figure 2A**). There were significant differences in the OD values for IgM-N between the severe and mild cases and between the critical and mild cases at 11–21 days after onset (severe vs. mild, *p* < 0.001; critical vs. mild, *p* = 0.020) (**Figure 2A**). The OD value for IgM-S1 was higher in the mild cases than in the severe and critical cases within 10 days after onset (critical vs. mild, *p* = 0.007) (**Figure 2B**). At 11–21 days after onset, the OD value for IgM-S1 was still significantly higher in the mild cases than in the critical cases (critical vs. mild, *p* = 0.007) (**Figure 2B**), and this difference disappeared at 22 days after onset (critical vs. mild, *p* = 0.065) (**Figure 2B**). There were significant differences in the OD values for IgG-N (severe vs. mild, *p* < 0.001; critical vs. mild, *p* < 0.001; critical vs. severe, *p* = 0.012) and IgG-S1 (severe vs. mild, *p* = 0.004; critical vs. mild, *p* < 0.001; critical vs. severe, *p* = 0.002) among the 3 groups at 11–21 days after onset, and the difference between the severe and critical cases disappeared at 22 days after onset (critical vs. severe, *p* = 0.069) (**Figure 2C and 2D**).

**Figure 2.**
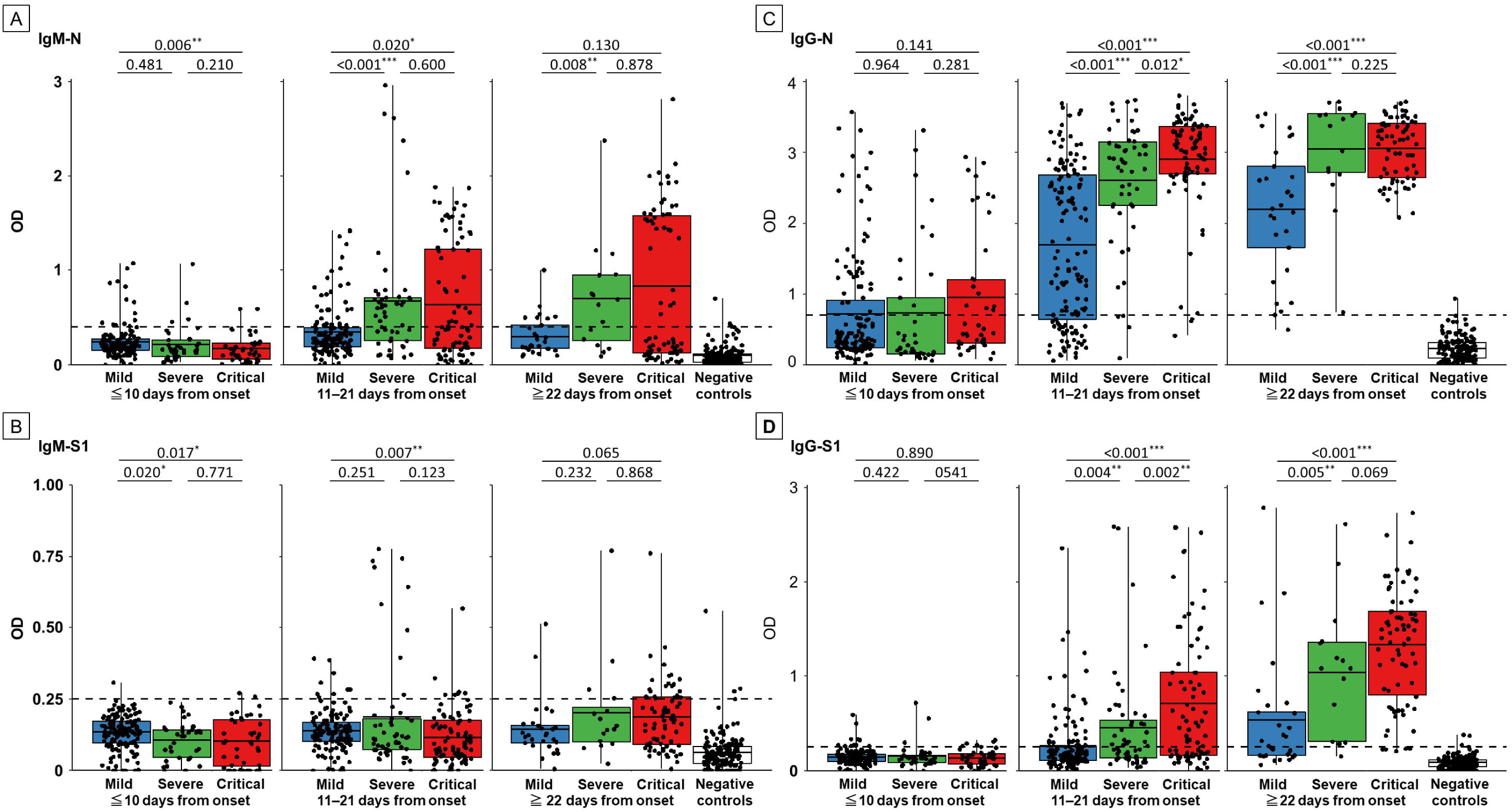
Comparison between disease severity and the antibody response for each ELISA. (A) IgM-N, (B) IgM-S1, (C) IgG-N, and (D) IgG-S1 in serum specimens collected at different time points from symptom onset. Blue box, mild cases; green box, severe cases; red box, critical cases; and white box, negative controls. Black plots indicate the OD value of each serum specimen. Black horizontal lines indicate the optimal cut-off values for each ELISA based on the results of 150 negative controls. The correlation coefficient was calculated using Wilcoxon’s rank-sum test. \**p* < 0.05, \*\**p* < 0.01, and \*\*\**p* < 0.001.

### Two antibody response patterns in severe and critical cases

A strong IgM-N response pattern (OD > 1.5) was observed in only the severe and critical cases during the clinical course (**Figures 1A and 2A**). A strong IgM-N pattern (OD > 1.5) was observed in 6.5% (2/31) of the severe cases and in 35.0% (7/30) of the critical cases. The 2 types of longitudinal antibody response among samples from individual patients in the severe and critical cases are shown in **Figure 3**: a strong IgM-N pattern (**Figure 3A–C**) and a weak IgM-N pattern (**Figure 3D–F**). In the strong IgM response pattern, IgM-N and IgG-N increased simultaneously. In both response patterns, the OD values of IgG-N increased earlier than those of IgG-S1 during the clinical course.

**Figure 3.**
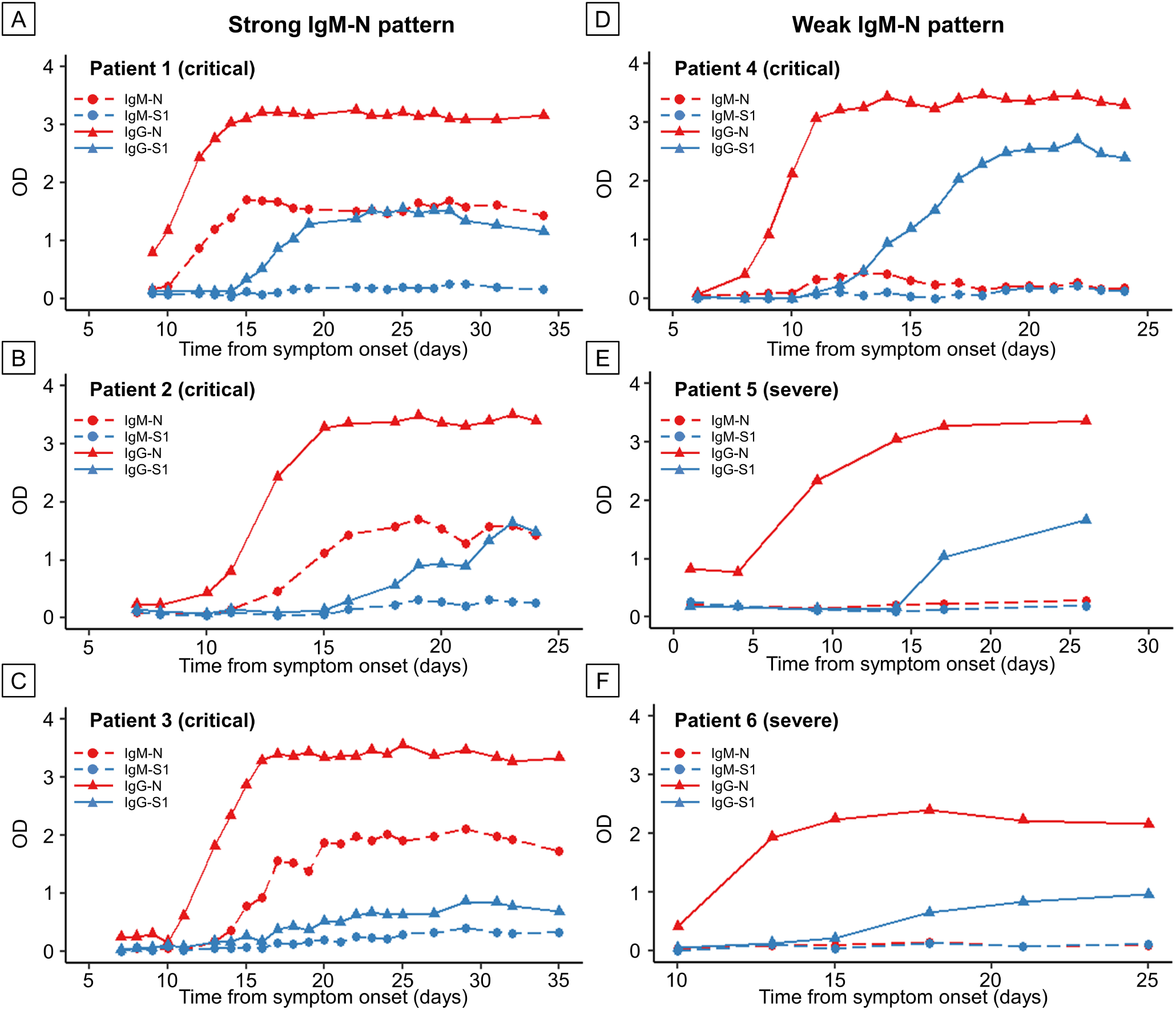
Antibody response patterns in severe and critical cases in 6 representative patients. Strong IgM-N patterns (A–C) and weak IgM-N patterns (D–F). Red, N protein; blue, S1 protein; dashed line and circle plots, IgM antibody assay; and solid line and triangle plots, IgG antibody.

### Seroconversion rate of antibodies in patients with COVID-19 in Japan

The seroconversion rates of each ELISA at the optimal cut-off value in the clinical course of these patients with COVID-19 are shown in **Figure 4**. Based on the results for 150 negative samples from patients without COVID-19, the optimal OD cutoff values were determined as 0.4 and 0.25 for IgM-N and IgM-S1, respectively, and 0.7 and 0.26 for IgG-N and IgG-S1, respectively (**Figure 2A-D**). Among all patients, the seroconversion rate of IgG was higher than that of IgM against both N and S1 during the clinical course (**Figure 4A and 4B**). In a comparison of the seroconversion rate according to disease severity, the seroconversion rate of IgG was also higher than that of IgM against both N and S1 (**Figure 4–H**). Except for 1 mild case, IgG-N seroconversion was observed in all 3 groups during the clinical course (mild, 95.7%; severe, 100.0%; critical, 100.0%). On the other hand, the seroconversion rates during the clinical course were higher in the severe and critical cases than in the mild cases for IgM-N (mild, 30.4%; severe, 70.8%; critical, 61.1%), IgM-S1 (mild, 8.7%; severe, 44.4%; critical, 44.4%), and IgG-S1 (mild, 47.8%; severe, 88.9%; critical, 94.4%) **(Figure 4C–H)**.

**Figure 4.**
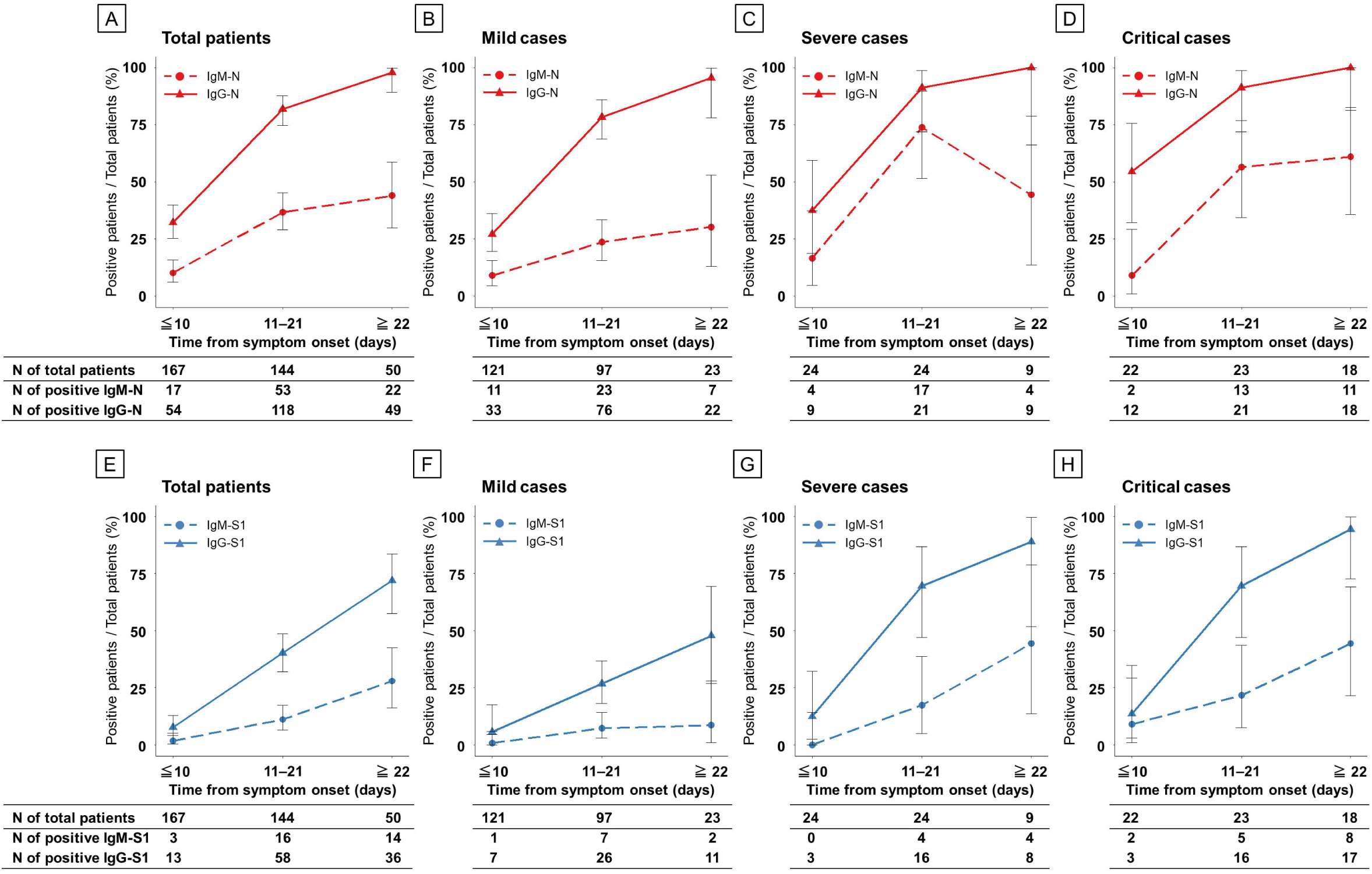
Seroconversion rate of ELISAs in patients with COVID-19 according to disease severity during the clinical course. Antibodies against N protein (A–D) and S1 protein (E–H). Red, N protein; blue, S1 protein; dashed line, IgM antibody assay; and solid line, IgG antibody.

## Discussion

Here, we presented analytical results for antibody response patterns according to disease severity in patients with COVID-19 in Japan. We showed several important features of the antibody response patterns in these patients. First, IgM-N and IgG-N increased at the same time, and the peak OD values for IgM-N and IgG-N appeared simultaneously. In addition, we showed that the seroconversion rate of IgG antibodies was higher than that of IgM antibodies, even in the early phase after symptom onset (<10 days). In viral infections such as Dengue virus that can cause reinfection, IgM becomes detectable earlier than IgG after the first viral infection. In the second infection, IgG is detectable earlier than or around the same time as IgM, and the titer of IgG increases rapidly after infection, while the titer of IgM becomes lower than during the first viral infection [17]. Sun et al. and Wang et al. reported the same antibody response patterns as described here in Chinese patients with COVID-19, and suggested they were affected by potential cross-reactivity of the humoral response between SARS-CoV-2 and other human coronaviruses (HCoVs) [4, 10]. In Japan, no outbreaks of SARS-CoV or Middle East respiratory syndrome-CoV have been reported; thus, cross-reactivity may have been caused by past infection with other seasonal HCoVs, such as HCoV-229E, HCoV-OC43, HCoV-NL63, and HCoV-HKU1. Although seroprevalence data for seasonal HCoVs in the Japanese population are not available, seasonal epidemics of HCoVs occur mainly during the winter in Japan [18]. Thus, the seroprevalence rate of seasonal HCoVs is expected to be high in the Japanese population. The antibody response patterns in our patient population support the hypothesis that cross-reactivity occurred in patients with COVID-19 in Japan. In addition, the low production of IgM antibody against N and S1 in our population can explain the low sensitivity of serological tests for IgM antibodies in Japan [19]. Thus, it is necessary to pay attention to the false-negative results of serological tests used for the initial diagnosis of COVID-19 in the clinical setting.

Second, a stronger IgM-N, IgG-N, and IgG-S1 response was observed in the severe and critical cases than in the mild cases of COVID-19, as observed in previous studies [4, 7, 10]. Several viral infections, such as Dengue virus, Ebola virus, and SARS-CoV, can cause antibody-dependent enhancement (ADE), which is the phenomenon by which antibodies paradoxically provide a means of enhancing virus entry and replication. A high viral load can amplify the secretion of cytokines by virus-infected cells and T cells and cause a cytokine storm, which consequently leads to increased viral pathogenicity and disease severity [7, 20, 21]. As in other viral infections, researchers have assumed that ADE may be one of the factors responsible for disease exacerbation in COVID-19 [7, 22]. Interestingly, a strong IgM-N response was observed in only the severe and critical cases. In patients infected with Ebola virus, it has been reported that IgM antibodies against viral antigens are mainly associated with ADE, and ADE may facilitate the rapid spread of the virus in the host during the early phase of infection [21]. Further studies are warranted to determine whether the excess production of antibodies is induced by only severe disease or if it is partly responsible for disease exacerbation in patients with COVID-19.

Third, a low seroconversion rate and weak response for IgG-S1 were observed in the mild cases. In our study, seroconversion of IgG-S1 antibody was observed in 47.8% of the mild cases, while it was detected in approximately 90% of the severe and critical cases. Several reports have also shown correlations between IgG-S1, RBD, and neutralizing antibody levels with disease severity, and a relatively low level of antibodies is observed in mild cases [23, 24]. It is assumed that SARS-CoV-2 infection is well controlled by the innate immune response and cellular immunity in mild cases. Additionally, Long et al. and Ibarrondo et al. showed the time course of a decrease in neutralizing antibodies at 2–3 months after initial infection in mild and asymptomatic cases [25, 26]. Taken together, patients who are asymptomatic and mild cases may have low levels of antibodies and a short period of humoral immunity.

This study has some limitations. First, we used only commercial ELISAs to detect the antibodies; thus, it is possible that the antibody response patterns are assay-specific. There is a possibility that the absence of seroconversion in several patients was caused by the test conditions and antigens used in the ELISAs utilized in our study. Second, cross-reactivity between SARS-CoV-2 and seasonal HCoVs in COVID-19 was suspected, but it was not examined in this study. Further investigations are required to verify the antibody response patterns by using several different serological methods and to examine cross-reactivity between SARS-CoV-2 and seasonal HCoVs.

## Conclusion

The findings of this study showed a stronger antibody response in Japanese patients with more severe COVID-19 and a low seroconversion rate of IgG-S1 in mild cases after infection. In addition, the antibody response patterns in the Japanese population suggest that there is cross-reactivity between SARS-CoV-2 and past infection with other HCoVs.

## Data Availability

The raw data used in this study is available from the corresponding author upon reasonable request.

## Acknowledgments

We thank the clinical laboratory technicians at the Self-Defense Forces Central Hospital for sample collection, and all members of the COVID-19 Task Force at the Self-Defense Forces Central Hospital and participating members drawn from other institutes of the Japan Self-Defense Force.

## References

[1] Huang C, Wang Y, Li X, Ren L, Zhao J, Hu Y, et al. Clinical features of patients infected with 2019 novel coronavirus in Wuhan, China. The Lancet. 2020;395:497–506.

[2] WHO. https://www.who.int/emergencies/diseases/novel-coronavirus-2019. [Latest accessed 7/Aug/2020].

[3] Wu Z, McGoogan JM. Characteristics of and Important Lessons From the Coronavirus Disease 2019 (COVID-19) Outbreak in China: Summary of a Report of 72lJ314 Cases From the Chinese Center for Disease Control and Prevention. JAMA. 2020;323:1239–42.

[4] Sun B, Feng Y, Mo X, Zheng P, Wang Q, Li P, et al. Kinetics of SARS-CoV-2 specific IgM and IgG responses in COVID-19 patients. Emerg Microbes Infect. 2020;9:940–8.

[5] Kissler SM, Tedijanto C, Goldstein E, Grad YH, Lipsitch M. Projecting the transmission dynamics of SARS-CoV-2 through the postpandemic period. Science. 2020;368:860–8.

[6] Shen L, Wang C, Zhao J, Tang X, Shen Y, Lu M, et al. Delayed specific IgM antibody responses observed among COVID-19 patients with severe progression. Emerg Microbes Infect. 2020;9:1096–101.

[7] Huang AT, Garcia-Carreras B, Hitchings MDT, Yang B, Katzelnick L, Rattigan SM, et al. A systematic review of antibody mediated immunity to coronaviruses: antibody kinetics, correlates of protection, and association of antibody responses with severity of disease. medRxiv. 2020:2020.04.14.20065771.

[8] Ju B, Zhang Q, Ge J, Wang R, Sun J, Ge X, et al. Human neutralizing antibodies elicited by SARS-CoV-2 infection. Nature. 2020. DOI:10.1038/s41586-020-2380-z.

[9] Brochot E, Demey B, Touze A, Belouzard S, Dubuisson J, Schmit J-L, et al. Anti-Spike, anti-Nucleocapsid and neutralizing antibodies in SARS-CoV-2 inpatients and asymptomatic carriers. medRxiv. 2020:2020.05.12.20098236.

[10] Wang Y, Zhang L, Sang L, Ye F, Ruan S, Zhong B, et al. Kinetics of viral load and antibody response in relation to COVID-19 severity. J Clin Invest. 2020. DOI:10.1172/jci138759.

[11] Walls AC, Park YJ, Tortorici MA, Wall A, McGuire AT, Veesler D. Structure, Function, and Antigenicity of the SARS-CoV-2 Spike Glycoprotein. Cell. 2020;181:281-92.e6.

[12] Okba NMA, Müller MA, Li W, Wang C, GeurtsvanKessel CH, Corman VM, et al. Severe Acute Respiratory Syndrome Coronavirus 2-Specific Antibody Responses in Coronavirus Disease 2019 Patients. Emerg Infect Dis. 2020;26:1478–1488.

[13] Premkumar L, Segovia-Chumbez B, Jadi R, Martinez DR, Raut R, Markmann A, et al. The receptor binding domain of the viral spike protein is an immunodominant and highly specific target of antibodies in SARS-CoV-2 patients. Sci Immunol. 2020;5:eabc8413.

[14] Amanat F, Stadlbauer D, Strohmeier S, Nguyen THO, Chromikova V, McMahon M, et al. A serological assay to detect SARS-CoV-2 seroconversion in humans. Nature Medicine. 2020. DOI:10.1038/s41591-020-0913-5.

[15] Chen X, Li R, Pan Z, Qian C, Yang Y, You R, et al. Human monoclonal antibodies block the binding of SARS-CoV-2 spike protein to angiotensin converting enzyme 2 receptor. Cellular & Molecular Immunology. 2020;17:647–9.

[16] Shirato K, Nao N, Katano H, Takayama I, Saito S, Kato F, et al. Development of Genetic Diagnostic Methods for Novel Coronavirus 2019 (nCoV-2019) in Japan. Jpn J Infect Dis. 2020. DOI:10.7883/yoken.JJID.2020.0611.

[17] Peeling RW, Artsob H, Pelegrino JL, Buchy P, Cardosa MJ, Devi S, et al. Evaluation of diagnostic tests: dengue. Nat Rev Microbiol. 2010;8:S30–8.

[18] Matoba Y, Abiko C, Ikeda T, Aoki Y, Suzuki Y, Yahagi K, et al. Detection of the human coronavirus 229E, HKU1, NL63, and OC43 between 2010 and 2013 in Yamagata, Jpn J Infect Dis. 2015;68:138–41.

[19] Imai K, Tabata S, Ikeda M, Noguchi S, Kitagawa Y, Matuoka M, et al. Clinical evaluation of an immunochromatographic IgM/IgG antibody assay and chest computed tomography for the diagnosis of COVID-19. J Clin Virol. 2020;128:104393.

[20] Katzelnick LC, Gresh L, Halloran ME, Mercado JC, Kuan G, Gordon A, et al. Antibody-dependent enhancement of severe dengue disease in humans. Science. 2017;358:929–32.

[21] Takada A, Ebihara H, Feldmann H, Geisbert TW, Kawaoka Y. Epitopes required for antibody-dependent enhancement of Ebola virus infection. Jpn J Infect Dis. 2007;196 Suppl 2:S347–56.

[22] French MA, Moodley Y. The role of SARS-CoV-2 antibodies in COVID-19: Healing in most, harm at times. Respirology. 2020;25:680–2.

[23] Liu L, To KK, Chan KH, Wong YC, Zhou R, Kwan KY, et al. High neutralizing antibody titer in intensive care unit patients with COVID-19. Emerg Microbes Infect. 2020;9:1664–70.

[24] Wu F, Wang A, Liu M, Wang Q, Chen J, Xia S, et al. Neutralizing antibody responses to SARS-CoV-2 in a COVID-19 recovered patient cohort and their implications. medRxiv. 2020:2020.03.30.20047365.

[25] Long Q-X, Tang X-J, Shi Q-L, Li Q, Deng H-J, Yuan J, et al. Clinical and immunological assessment of asymptomatic SARS-CoV-2 infections. Nature Medicine. 2020.

[26] Ibarrondo FJ, Fulcher JA, Goodman-Meza D, Elliott J, Hofmann C, Hausner MA, et al. Rapid Decay of Anti–SARS-CoV-2 Antibodies in Persons with Mild Covid-19. New England Journal of Medicine. 2020. DOI:10.1056/NEJMc2025179.

